# Antidepressants are associated with decreased white blood cell counts beyond the effect of statins in individuals with co-morbid depression and coronary artery disease: A longitudinal analysis in the Vanderbilt University Medical Center and All of Us cohorts

**DOI:** 10.1101/2025.10.13.25337946

**Authors:** Kritika Singh, Freida A. Blostein, Quinn S. Wells, Emily C. Hodges, Nancy J. Cox, Lea K. Davis

## Abstract

Depression and coronary artery disease (CAD) are comorbid, prevalent conditions responsible for significant morbidity and mortality. Understanding the co-occurrence of these conditions can facilitate early diagnosis and treatment recommendations. Routinely prescribed treatments are available for both CAD and depression, and interactions between serotonin signaling, the immune system, and cardiovascular system suggest cross-treatment of CAD and depression may be indicated. In this study we use repeated, longitudinal WBC count measures from over one million participants in the Vanderbilt University Medical Center (VUMC) electronic health records (EHR) to characterize changes in systemic inflammation in response to CAD and depression diagnoses and their respective treatments. After controlling for potential confounders, individuals with comorbid depression-CAD have higher median WBC counts than controls and individuals with depression only but not higher than individuals with CAD only. In our longitudinal modeling cohort of individuals with both depression and CAD, there was a significant decrease in WBC count after statin initiation and after anti-depressant initiation. Additionally, initiating antidepressants after statins was associated with a further decrease in WBC count, however the reverse was not true. This work highlights the continued importance and necessity of evaluating the immunomodulatory effects of depression and CAD and their respective treatments in individuals with both depression and CAD.

## Introduction

Depression and coronary artery disease (CAD) are prevalent conditions responsible for significant morbidity and mortality^1,2^. Depression and CAD are also highly comorbid: approximately 40% of individuals who have one diagnosis will develop the other^3^. Understanding the co-occurrence of these conditions can facilitate early diagnosis and treatment recommendations.

Both CAD and depression exhibit inflammatory etiologies^4–7^. Acute inflammation is characterized by short-term, abnormally high levels of pro-inflammatory biomarkers circulating in the blood, typically in response to injury or infection^8^. Elevated but not abnormal levels of pro-inflammatory markers can be indicative of chronic inflammation. CAD, formerly considered a lipid accumulation-mediated disease, also involves an ongoing, pro-inflammatory response with phases of both acute and chronic inflammation^5,7,9–11^. As CAD progresses, pro-inflammatory biomarkers such as C-Reactive protein (CRP), white blood cells (WBCs, i.e., leukocytes and monocytes) and inflammation-associated prothrombotic markers including platelets, increase as compared to healthy controls^5,7,9–11^. Similarly, the onset and severity of depression is associated with chronic inflammation, including increases in leukocytes, CRP, tumor necrosis factor-alpha (TNF-α), interleukin-1β (IL-1β) and IL-6 in brain and peripheral blood samples of individuals with depression as compared to healthy controls^4,6,12–14^.

However, unlike CAD, instances of acute inflammation are not classically associated with depression. While both CAD and depression exhibit inflammatory etiologies, none of these studies have simultaneously described their joint and independent relationship with inflammatory markers.

Routinely prescribed therapeutic treatments are available for both CAD and depression, and interactions between serotonin signaling, the immune system, and cardiovascular system suggest cross-treatment of CAD and depression may be indicated^15–18^. Statins are a first-line therapeutic treatment for dyslipidemia, an integral precursor to atherosclerosis^19,20^. Statins also have anti-inflammatory properties ^21–23^. This may explain why the use of statins was associated with significant reduction in the risk of depression diagnosis in individuals who have had a cardiac event^24,25^.

Antidepressants are first-line pharmaceutical treatments for depression and are among the most prescribed medications in the United States, with the selective serotonin inhibitors (SSRIs) being the most commonly prescribed^26^. SSRIs modulate uptake of serotonin but also suppress serum and plasma levels of pro-inflammatory mediators in patients with major depression ^27–29^. Past research indicates that SSRI’s were associated with a reduced risk of arrhythmia and myocardial infarction^30,31^. This evidence suggests cross-prescribing of antidepressants and statins could benefit patients; however, it is not yet clear whether statins and antidepressants have independent, overlapping, or synergistic effects on inflammation^30,31^.

Neuroinflammation and peripheral inflammation are hypothesized to play an important role in both depression and CAD respectively, providing a potential common biological pathway that may link neuroinflammation in depression together with atherosclerotic inflammation in CAD. A robust body of literature points to the role of inflammation in both depression and CAD independently, but few studies have investigated their joint effects^8,16,31^. In this study we use repeated, longitudinal WBC count measures from over one million participants in the Vanderbilt University Medical Center (VUMC) electronic health records (EHR) to characterize changes in systemic inflammation in response to CAD and depression diagnoses and their respective treatments.

## Methods

### Study populations: VUMC EHR and All of Us

VUMC is a tertiary care center that provides inpatient and outpatient care in Nashville, TN. The VUMC EHR was established in 1990 and includes data on billing codes from the International Classification of Diseases, 9th and 10th editions (ICD-9 and ICD-10), Current Procedural Terminology codes, medications, laboratory values, reports, and clinical documentation. This analysis uses the de-identified mirror of the EHR, which includes over 3.2 million patient records. In the deidentification, dates are shifted to maintain privacy but the order of events and time between them are preserved, enabling longitudinal investigations. The VUMC Institutional Review Board oversees BioVU and approved this project (IRB#172020, 201203).

To be eligible for inclusion in the VUMC cohort, participants needed to meet a simple “medical home” heuristic of having at least five medical codes on different days over a period of at least three years. This inclusion criteria reduces the missingness of clinical data and enriches the sample for patients who receive their primary care at VUMC. Participants were also required to have at least one clinical measurement of WBC in an out-patient setting (details below). Participants with conditions known to impact WBC counts including cancer diagnosis or autoimmune disease phecodes (Type 1 diabetes – 250.1, ulcerative colitis – 555.21, rheumatoid arthritis - 714, and lupus - 695.41) were excluded. Phecodes are a higher order combination of at least two related ICD codes occurring on two different days and were curated using the R PheWAS package^32^

We replicated all analyses using EHR and health questionnaire data from All of Us, a large-scale biobank consisting of comprehensive health data from participants reflecting the diversity of the United States of America (Supplemental Methods).

### Outcome variable curation: WBC count measurements

WBC counts were extracted and cleaned from the VUMC EHR using QualityLabs, a previously described laboratory-wide cleaning and QC pipeline^33^. To avoid bias associated with acute illness, injury, and corresponding treatments during emergency visits or in-patient hospitalizations, we included only WBC counts from out-patient visit dates. All non-numeric values were removed, laboratory values were required to be recorded in the same unit, and values greater than four standard deviations from the sample mean were removed. To facilitate interpretation in statistical modeling, all outpatient associated WBC measures were normalized using a z-score transformation so that effect estimates could be interpreted per one standard deviation increase in WBC count. In cross-sectional analyses, a per-person summary value was created by taking the median value across WBC measurements.

The same pipeline was applied in All of Us. However, we included measurements from visits missing a visit type designation because these represented a significant fraction of WBC measurements in All of Us (65% of measurements).

### Exposure variable definitions: Clinical diagnoses and medication initiation

We defined cases and controls in the VUMC EHR using phecodes. Participants with a phecode for *both* depression (296.2, which includes major depressive disorder) and CAD (411.4) were considered our exposed group or cases (herewith abbreviated (m)dCAD). We considered four comparison groups: 1) unscreened controls (any non-case participants which also includes those with either CAD OR depression, but not both conditions), 2) screened controls (any participants with *neither* CAD *nor* depression phecodes), 3) depression only controls (participants with depression phecodes but not CAD phecodes) and 4) CAD only controls (participants with CAD phecodes but not depression phecodes). We identified the first diagnosis of CAD and/or depression based on the date of the earliest ICD9 or 10 code used to define the phecodes for CAD and/or depression (Supplemental Table 1).

Similarly, we identified participants who had any record of statins (Supplemental Table 2) and/or antidepressants in the VUMC EHR (as described elsewhere^34^) and extracted the date at earliest record as the medication initiation date.

In All of Us, we used similar definitions for clinical diagnoses and medication initiation (Supplemental Methods).

### Other covariates

We identified potential confounders for associations between depression, CAD, and WBC counts based on a literature review. These included EHR-defined race, age, sex, BMI, and diagnoses for tobacco use disorder, type 2 diabetes, hypertension, length of medical record, and number of WBC measurements per-person. Tobacco use disorder, type 2 diabetes and hypertension definitions were derived from phecodes, and the other variables were directly sourced from the EHR (Supplemental Methods).

To verify our results, we identified positive and negative control conditions for use in multivariable modeling. We identified two diseases (Type-2 diabetes and ulcerative colitis) with known inflammatory biology as positive controls and four diseases (substance use disorder, glaucoma, migraine, and osteoarthritis) that are not known to include inflammatory biology as negative controls, based on prior literature^35–39^. The conditions were also defined using phecodes, and the first date was extracted based on the first date of the ICD codes used to define the phecodes (Supplemental Table 1).

### Statistical analyses

To test if participants with (m)dCAD have higher WBC counts on average than other patients, we performed parallel linear regression models comparing per-person median WBC counts in cases versus each control definition separately, while controlling for race, sex, age, median BMI, type 2 diabetes, hypertension, tobacco use disorder, number of WBC counts, the length of the medical record, and an indicator variable for “ever record of antidepressants”.

To test if WBC counts changed over time in response to medication initiation, we ran parallel mixed effects linear regression models using the REML R package. We created time varying covariates (TVCs) for statin and antidepressant initiation (not medicated/medicated), defined according to whether the WBC measurement date preceded or succeeded the first medication date. We again performed these regressions in three mutually exclusive subsets of the analytic sample: 1) depression only controls, 2) CAD only controls, and 3) comorbid (m)dCAD cases, including as the main fixed effect predictor respectively an antidepressant TVC, a statin TVC, or both an antidepressant and statin TVC. To test if WBC counts changed in response to initial diagnosis, we performed similar regressions for depression and CAD diagnosis status, using the same TVCs based on whether the WBC measurement date preceded or succeeded the first diagnosis date.

For patients who were prescribed both antidepressants and statins, we tested whether order of medication initiation had an impact on WBC trajectories during treatment. To address this question we ran mixed effects linear regression models in two subsets: 1) participants with antidepressant initiation prior to statin initiation, and 2) participants with antidepressant initiation subsequent to statin initiation. These models included all participants with both statin and antidepressant records, regardless of case-control status. Each regression included an antidepressant and statin TVC.

In each mixed effects model described above we included a random intercept for participant, a random slope for age at WBC measurement, and fixed effects for race, age, sex, median BMI, tobacco use disorder, type 2 diabetes and number of WBC measurements. We applied Bonferroni correction for the 18 independent tests conducted during multivariable regression i.e. α = 0.05/18 = 0.0027.

## Results

### Individuals with (m)dCAD have higher median WBC counts than individuals with only depression or only CAD

After applying inclusion and exclusion criteria, the VUMC cohort consisted of 276,586 screened controls, 32,185 participants with depression only, 22,007 individuals with CAD only, and 3,603 individuals with comorbid (m)dCAD. Individuals with (m)dCAD had the highest normalized median WBC counts of all participants (Table 1). Median WBC counts among those with (m)dCAD were on average 0.06 SD higher than those with CAD only (Welch’s two sample t-test *p*=6e-5), 0.07 SD higher than those with depression only (*p=*2e-6) and 0.05 SD higher than screened controls (*p*=6e-5, Figure 1). Individuals with (m)dCAD were more likely than other participants - including participants with only depression or only CAD - to have other chronic conditions, such as tobacco use disorder, hypertension and type 2 diabetes (Table 1). All of Us participants (n=54,003 screened controls, n=21,865 with depression only, n=5,995 with CAD only, and n=3,796 with (m)dCAD) were less likely to have an EHR reported race of white and more likely to have Type 2 Diabetes than VUMC participants. (Supplemental Table 3).

**Figure 1:**
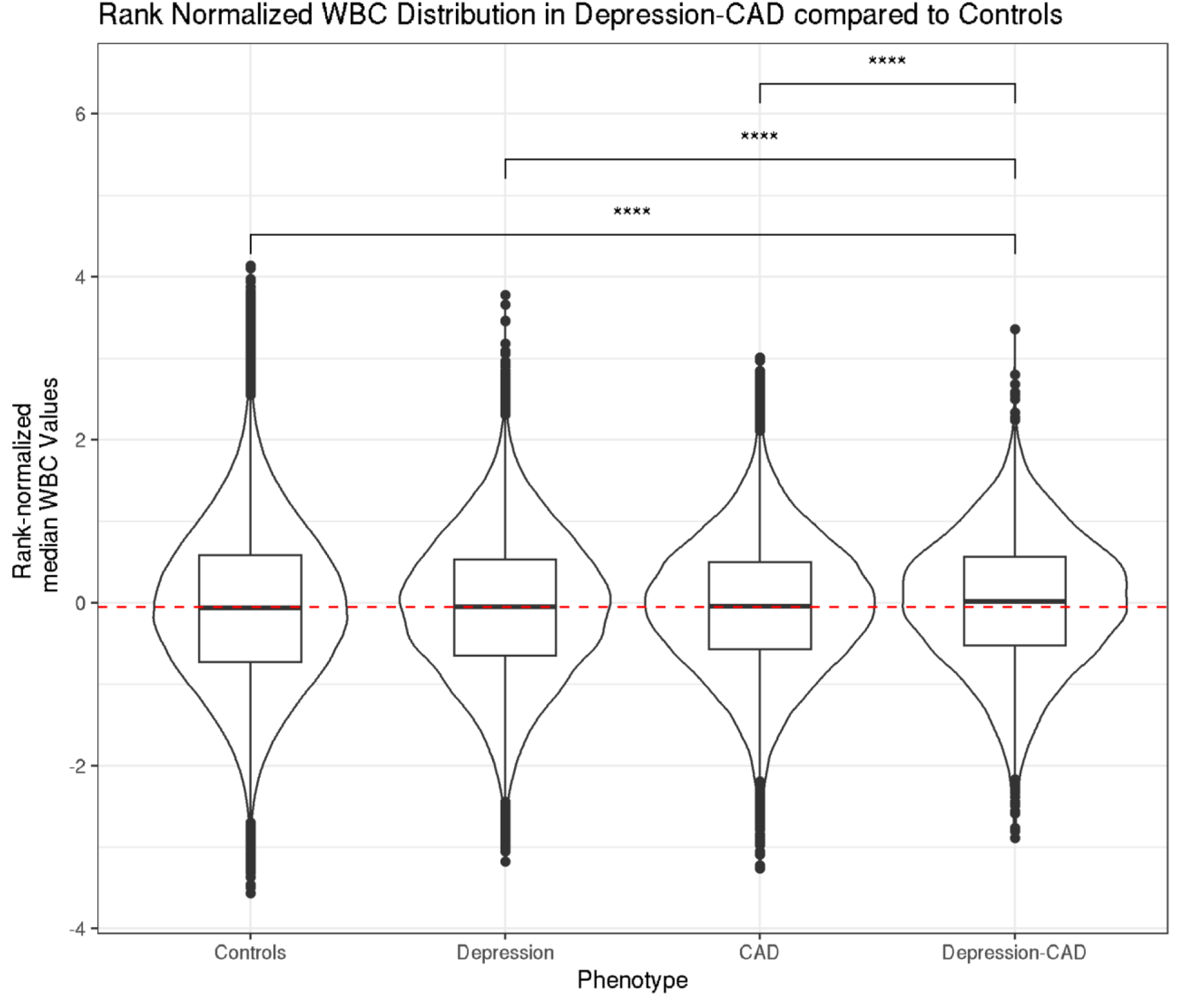
Distribution of inverse rank normalized WBC per-person medians among those with comorbid depression and coronary artery disease (CAD), depression only, CAD only, and without either condition among 334,294 participants in the Vanderbilt University Medical Center Electronic Health Record.

**Table 1:** White blood cell counts, demographic characteristics and medical conditions among 334,381participants in the Vanderbilt University Medical Center Electronic Health Record.

| **Characteristic** | **Controls,**<br>**N = 276,586<sup>i</sup>** | **Depression,**<br>**N = 32,185<sup>i</sup>** | **CAD,**<br>**N = 22,007<sup>i</sup>** | **(m)dCAD,**<br>**N = 3,603<sup>i</sup>** |
| --- | --- | --- | --- | --- |
| Per-person WBC count<br>medians (thousand per<br>microliter) | 7.30 (5.90, 9.10) | 7.30 (6.05, 8.90) | 7.35 (6.20, 8.80) | 7.50 (6.30, 9.00) |
| Rank-normalized per-person<br>WBC count medians | -0.06 (-0.73, 0.60) | -0.06 (-0.65, 0.53) | -0.04 (-0.59, 0.50) | 0.02 (-0.52, 0.56) |
| Medical record length (days)<br>(mean (sd)) | 4,086 (2,117) | 4,690 (2,244) | 4,150 (2,014) | 4,794 (2,235) |
| Number of WBC<br>measurements (mean (sd)) | 4 (8) | 7 (12) | 7 (10) | 15 (21) |
| EHR sex |  |  |  |  |
| Female | 168,338 (61%) | 22,884 (71%) | 7,156 (33%) | 1,680 (47%) |
| EHR race |  |  |  |  |
| White | 214,251 (77%) | 26,707 (83%) | 19,476 (88%) | 3,151 (87%) |
| Black | 40,250 (15%) | 4,069 (13%) | 1,912 (8.7%) | 385 (11%) |
| Asian | 7,193 (2.6%) | 416 (1.3%) | 215 (1.0%) | 18 (0.5%) |
| Other | 3,244 (1.2%) | 201 (0.6%) | 72 (0.3%) | 6 (0.2%) |
| Unreported | 8,766 (3.2%) | 453 (1.4%) | 181 (0.8%) | 5 (0.1%) |
| Multiple race categories | 2,254 (0.8%) | 284 (0.9%) | 122 (0.6%) | 34 (0.9%) |
| Alaskan native/Native<br>American | 628 (0.2%) | 55 (0.2%) | 29 (0.1%) | 4 (0.1%) |
| Median age across medical<br>record (years) | 34 (18, 52) | 40 (25, 55) | 66 (58, 74) | 63 (55, 72) |
| Median BMI across BMI measurements | 26 (21, 31) | 28 (23, 33) | 29 (26, 33) | 29 (25, 34) |
| Hypertension | 57,702 (21%) | 11,576 (36%) | 18,773 (85%) | 3,325 (92%) |
| Type 2 Diabetes | 18,184 (6.6%) | 3,822 (12%) | 7,291 (33%) | 1,555 (43%) |
| Tobacco use disorder | 11,703 (4.2%) | 5,111 (16%) | 4,131 (19%) | 1,359 (38%) |
| Ever antidepressants reported | 68,947 (25%) | 22,878 (71%) | 8,243 (37%) | 3,068 (85%) |
| Ever statins reported | 52,520 (19%) | 8,525 (26%) | 19,842 (90%) | 3,213 (89%) |
| Time between first statin report and first CAD ICD (days)<sup>2</sup> |  |  | 0 (-439, 1) | -11 (-1,121, 1) |
| Never statins |  |  | 2,165 (10%) | 390 (11%) |
| Time between first antidepressant report and first depression ICD (days)<sup>2</sup> |  | -112 (-1,126, 30) |  | -342 (-1,547, 7) |
| Never antidepressants |  | 9,307 (29%) |  | 535 (15%) |

<sup>1</sup> Median (IQR); Mean (sd) where indicated; n (%)
<sup>2</sup> Negative numbers indicate number of days prior to first ICD code that medication was initiated, positive numbers indicate number of days after first ICD code that medication was initiated
WBC = White blood cells; ICD=International classification of disease code; CAD=Coronary artery disease;
EHR=electronic health record; BMI=body mass index

After accounting for confounders in linear regression models, participants from VUMC with (m)dCAD had 0.18 standard deviations higher median WBC counts than screened controls (*P*=2e–30) and 0.14 standard deviations higher median WBC counts than participants with depression only (*P*=9e-17) (Figure 2, Supplemental Table 4). However, median WBC counts were similar between cases and participants with CAD only (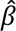=0.02, *P*=0.11). Results were similar in All of Us (Supplemental Table 4).

**Figure 2:**
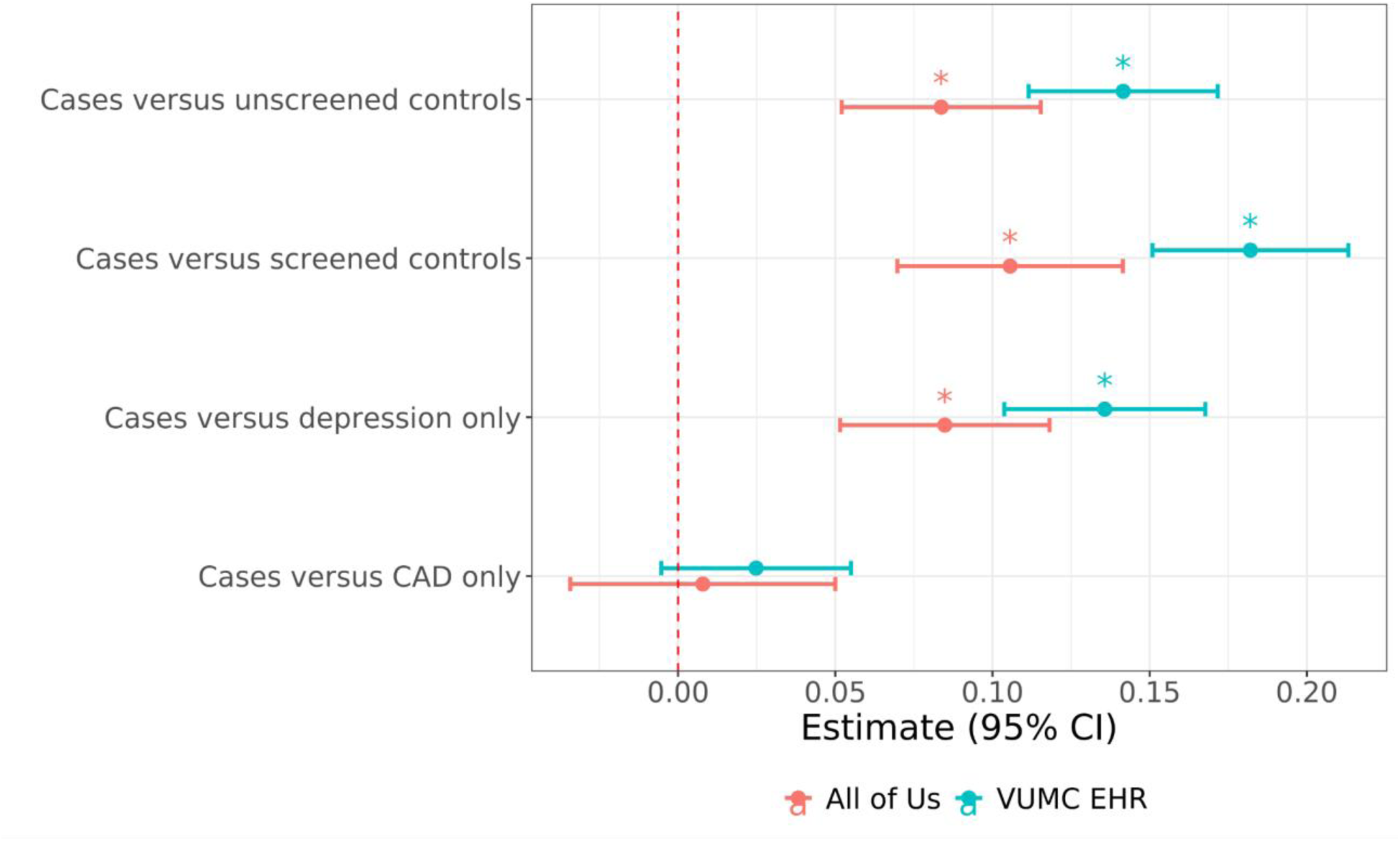
Effect estimates for difference in average median white blood cell (WBC) counts between (m)dCAD cases and four comparison groups in the Vanderbilt University Medical Center (VUMC) Electronic Health Record (EHR) cohort (blue) and All of Us cohort (red). Unscreened controls refers to participants without (m)dCAD, screened controls refers to participants without (m)dCAD, depression, or CAD. Effect estimates from multivariable regression models controlling for median age across medical record, sex, EHR-reported race, medical record length, number of white blood cell measurements, median BMI across record, any hypertension, any type 2 diabetes, and tobacco use disorder, and ever record of antidepressant.

### While both statins and antidepressants are associated with decreases in WBC counts, statins do not further reduce WBC counts when co-administered with antidepressants among those with (m)dCAD

We examined the effect of the first record of statin and antidepressant medications on WBC counts. Among participants with CAD only who received a statin (n=19,842), the statin TVC was associated with a 0.075 standard deviation decrease (*P*=2e-24) in WBC counts in longitudinal mixed effects models, after controlling for race, sex, age at WBC measurement (fixed effect and random slope), median BMI across record, hypertension, tobacco use disorder, type 2 diabetes, and an indicator variable for “ever antidepressant use”. Similarly, among participants with depression only who received antidepressants (n=22,878), the antidepressant TVC was associated with a 0.02 standard deviation decrease (*P*=2e-5) in WBC counts, after controlling for the same variables except substituting an indicator variable for “ever statins use” instead of “ever antidepressant use”. However, among individuals with (m)dCAD who received *both* antidepressants and statins (n=2,801), the antidepressant TVC was associated with a 0.05 standard deviation decrease in WBC (*P*=2e-4), but the statin TVC was not statistically significant after correcting for multiple testing (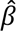=0.03, *P*=0.05, Figure 3 and 4). These results were replicated in All of Us (Table 2).

**Figure 3:**
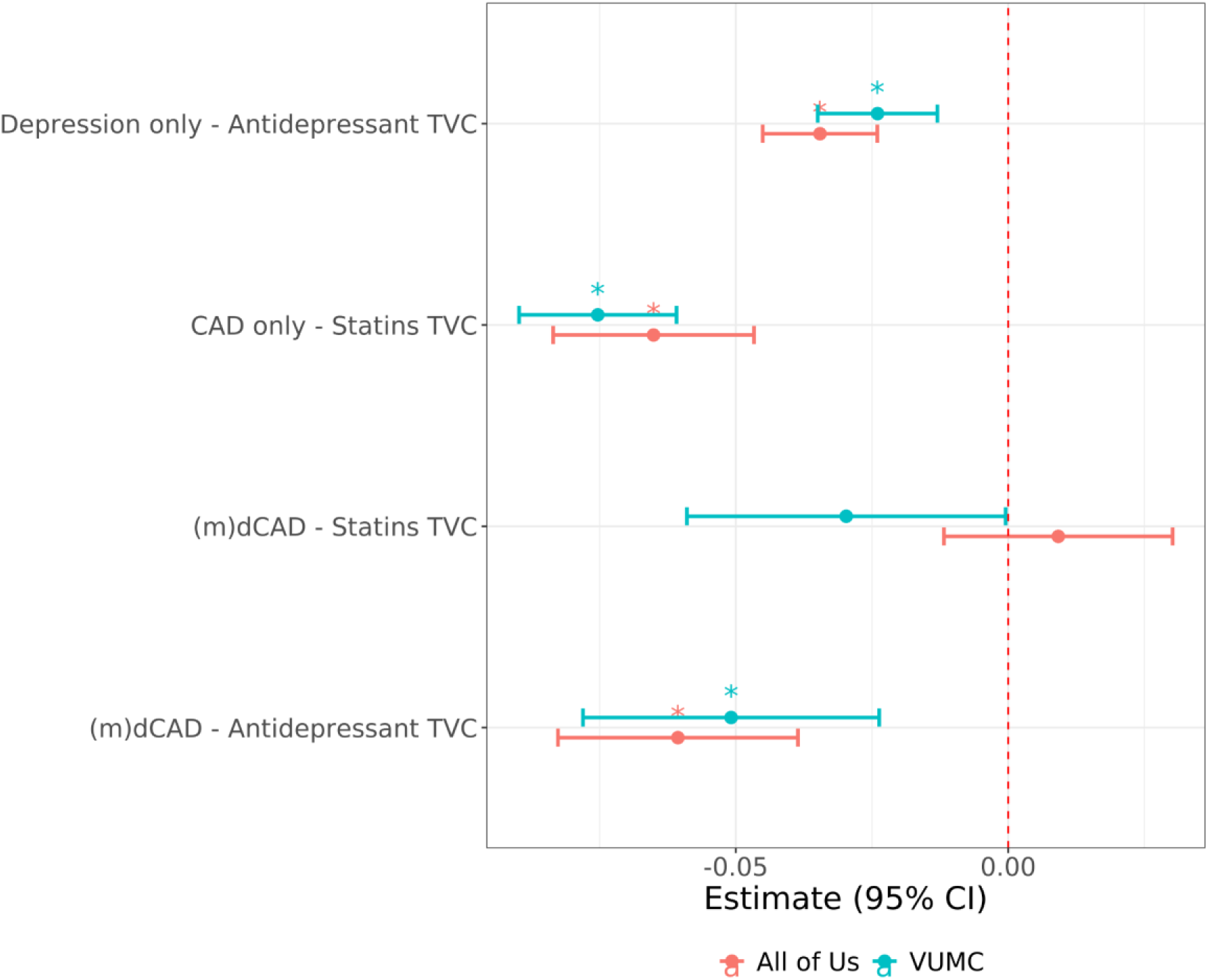
Effect estimates of average longitudinal change in white blood cell (WBC) counts by initiation of statins or antidepressants within three population subsets: those with depression only, those with CAD only, and those with (m)dCAD among participants from VUMC (blue) and All of Us (red). Effect estimates are fixed effect estimates for time varying covariates (TVCs) representing initiation of the medication, from multivariable mixed effect regression models controlling for age at WBC measurement, sex, EHR-reported race, medical record length, number of white blood cell measurements, median BMI across record, any hypertension, any type 2 diabetes, and tobacco use disorder. Among individuals with depression only, an additional covariate controlled for ever record of statins. Among individuals with CAD only, an additional covariate controlled for ever record of antidepressants. Models among those with (m)dCAD included TVCs for both antidepressant and statin initiation.

**Figure 4:**
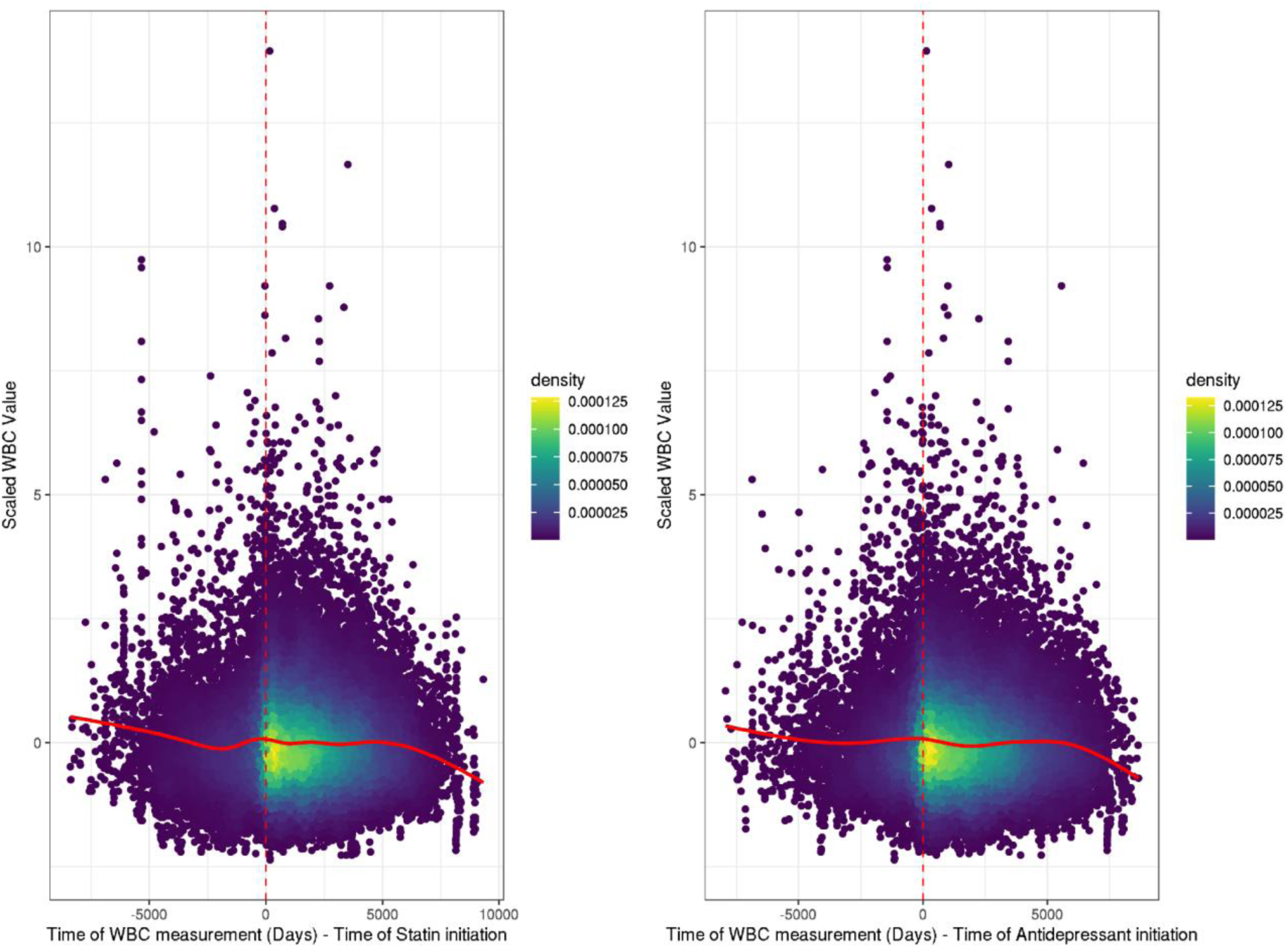
Density scatter plot with smoothed line showing white blood cell (WBC) values over time in relation to the first date of statin (red-dashed line, panel A) or antidepressant (red dashed line, panel B) among participants with (m)dCADi n the Vanderbilt University Medical Center Electronic Health Record cohort.

**Table 2:** Results from mixed effect linear regression models comparing the effect of first medication record (statins or antidepressants) on normalized white blood cell counts among individuals with depression only, coronary artery disease (CAD) only, and comorbid depression and CAD (m)dCAD in the Vanderbilt University Medical Center cohort and the All of Us cohort.

| Source | Cohort | Covariate | N | Estimate | Standard error | Lower 95% CI | Upper 95% CI | P value | Bonferroni significant |
| --- | --- | --- | --- | --- | --- | --- | --- | --- | --- |
| *Vanderbilt University Medical Center* |  |  |  |  |  |  |  |  |  |
|  | (m)dCAD | Antidepressant TVC | 2801 | -0.051 | 0.014 | -0.078 | -0.024 | 2.46e-04 | * |
|  | (m)dCAD | Statins TVC | 2801 | -0.030 | 0.015 | -0.059 | 0.000 | 4.63e-02 |  |
|  | CAD only | Statins TVC | 19842 | -0.075 | 0.007 | -0.090 | -0.061 | 0.00e+00 | * |
|  | Depression only | Antidepressant TVC | 22878 | -0.024 | 0.006 | -0.035 | -0.013 | 1.88e-05 | * |
| *All of Us* |  |  |  |  |  |  |  |  |  |
|  | (m)dCAD | Antidepressant TVC | 3796 | -0.061 | 0.011 | -0.083 | -0.039 | 1.00e-07 | * |
|  | (m)dCAD | Statins TVC | 3796 | 0.009 | 0.011 | -0.012 | 0.030 | 3.91e-01 |  |
|  | CAD only | Statins TVC | 5995 | -0.065 | 0.009 | -0.084 | -0.047 | 0.00e+00 | * |
|  | Depression only | Antidepressant TVC | 21865 | -0.035 | 0.005 | -0.045 | -0.024 | 0.00e+00 | * |

### First records of CAD and depression diagnosis codes are associated with decreases in WBC counts, and initial diagnosis is close in time to treatment initiation

The first record of a depression or CAD ICD code was also associated with a decrease in WBC counts in longitudinal mixed effect regression models in the VUMC cohort (Supplemental Table 5, Supplemental Figure 4). As expected in a medical setting, first records of depression and CAD diagnoses were frequently close in time to the first record of treatment for these conditions including antidepressants and statins, respectively (Supplemental Figures 1-3). In the VUMC cohort, the first statin record preceded the first CAD ICD code by 0 days or 11 days on average for those with CAD only or (m)dCAD, respectively, while first antidepressant record preceded depression ICD code by ∼4 or ∼11 months, on average for those with depression only or (m)dCAD (Table 1). Positive control conditions exhibited the same inverse association with WBC (Supplemental Figure 5), suggesting the first diagnosis in the VUMC EHR is a strong proxy for treatment initiation. In contrast, in the All of Us cohort, where statin report was less common and less likely to be concurrent with first CAD ICD code (Supplemental Tables 6-7), the first CAD ICD code was not associated with a decrease in WBC counts (Supplemental Table 5).

### Initiating antidepressants after statins is associated with further decreases in WBC counts, while initiating statins after antidepressants is not

Given the association between medications and declining WBC count, we investigated the impact of medication order on WBC count change over time among any participants with records of both medications (regardless of diagnosis). Among those with a first record of statins before the first record of antidepressants (n=15,000), the statin TVC was associated with a 0.08 standard deviation decrease in WBC counts (*P*< 2e-16), and the subsequent antidepressant TVC was associated with an additional 0.04 standard deviation decrease in WBC counts (*P*=6e-11). Among individuals with a first record of statins *after* the first record of antidepressants (n=16,426), the antidepressant TVC was associated with a 0.06 standard deviation decrease in WBC counts, while the subsequent statin TVC was not associated with a further decrease in WBC (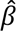=0.001, *P*=0.82, Supplemental Table 8, Supplemental Figures). Results for antidepressants were similar in All of Us. However, in All of Us the statin TVC was associated with an *increase* in WBC among those with antidepressant before statin initiation (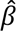= 0.03, *P*=0.0002) and was not associated with change in WBC among those with statins preceding antidepressant initiation (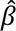= 0.003, *P*=0.77, Supplemental Table 8, Supplemental Results).

## Discussion

In two independent cohorts, we found that individuals with comorbid (m)dCAD had higher WBC counts than screened or unscreened controls. However, after controlling for potential confounders, individuals with (m)dCAD also had higher WBC counts than patients with depression only but similar WBC counts compared to patients with CAD only. Individuals with (m)dCAD had increased rates of comorbidities compared to screened and unscreened controls, including very high (>90%) rates of hypertension, which has previously been reported^39^. These findings suggest comorbid conditions such as tobacco use disorder, type 2 diabetes and hypertension, which are more common among those with (m)dCAD, account for some of the increase in inflammatory markers in those with (m)dCAD. In line with previous literature, our work supports the hypothesis that depression and CAD share an underlying inflammatory etiology, potentially contributing to the observed co-occurrence of these diseases^10,15,30,40^.

We advance on previous work by leveraging longitudinal data to investigate whether the timing of CAD and depression diagnosis and treatment influenced white blood cell count trajectories. We found that WBC counts decrease on average following the first record of depression and CAD diagnosis codes, likely because a clinical diagnosis in a healthcare setting is frequently linked to treatment initiation, and in fact treatment frequently precedes recorded diagnosis. We also investigated the effects of statin and antidepressant initiation on WBC count, first finding that antidepressant and statin initiation treatment each decreased WBC count measures in the depression only and CAD only patient populations, respectively. This is in line with previous work showing both statins and antidepressants have anti-inflammatory properties^21,23,28,41,42^.

Next, we demonstrate that antidepressants are associated with lower WBC counts in individuals with (m)dCAD independent of statin administration, in both the VUMC and All of Us cohorts. Furthermore, the independent association of antidepressant initiation with WBC count decline is not merely an artifact of medication temporal ordering. First antidepressant record was always associated with a decrease in WBC counts, both when antidepressants preceded statins and when statins preceded antidepressants. Conversely, the first statin record was not associated with further WBC declines among those for whom the first statin record followed their first antidepressant record. Associations between antidepressants and decreasing WBC count were consistent across the two cohorts, while the effect of statin initiation was weaker or reversed in All of Us, potentially reflecting different patterns of medication reporting and subsequent exposure misclassification in the two cohorts (Supplemental Results).

Previous studies have reported that antidepressants decrease the odds of subsequent myocardial infarctions in individuals with CAD^30,42–44^. Our work raises the hypothesis that the risk reduction may be at least partially mediated through an anti-inflammatory mechanism ^30,42^. Given the frequency of depression-CAD comorbidity, and our observation that antidepressants are associated with WBC declines even when administered after statins, there may be a clinical benefit to screening for depressive symptoms and initiating antidepressants in patients with known CAD. Since initiating statins after antidepressants was not associated with a further decrease in WBC counts, earlier initiation of antidepressants may yield greater anti-inflammatory benefits that can potentially improve cardiac related outcomes^42–43^.

Our work is one of the few studies to evaluate longitudinal changes in an inflammatory marker (WBC count) in individuals with both depression and/or CAD, in a large study population^4,38,44^. Additionally, we demonstrate the relationship between depression and CAD treatment i.e., statins and antidepressants and WBC count measures using EHR data in a longitudinal framework.

Despite its strengths, our study is limited by the well-established caveats of EHR based phenotyping which can suffer from clinician bias in diagnosis and ascertainment bias of lab traits. To mitigate this, we excluded individuals with acute illnesses such as autoimmune conditions and cancers from our analytic sample. We further excluded all WBC measures that were taken in an inpatient setting. In our results we observed that individuals with depression and CAD had significantly higher WBC counts than our selected controls. Our screened controls are from a tertiary healthcare system, so they may have other conditions that could elevate their WBC levels. As a result, healthier individuals from the general population could have lower WBC levels than the controls we used, which would bias our results towards the null. Secondly, linear mixed models assume linear effects in coefficients which may result in missing non-linear effects.

Thirdly, it was not possible to measure medication adherence, and this study did not include additional medications. Lastly, it is possible that WBC count reduction in response to statins, antidepressants and their combination, could be a dose-sensitive phenomenon, however, in this study we were unable to include dosage data ^38^.

Nonetheless, our work shows that there is a modest increase in WBC counts in (m)dCAD compared to conservatively defined controls. We additionally observe a decrease in WBC count levels after first antidepressant treatment in depression, CAD, and (m)dCAD. Lastly, our work adds to the literature by showing that initiating antidepressants after statins is associated with a *further* decrease in WBC count. Overall, this work highlights the continued importance and necessity of evaluating the immunomodulatory effects of depression and CAD and their respective treatments in individuals with both depression and CAD.

## Code and Data availability

All code to conduct these analyses is available at https://bitbucket.org/davislabteam/mdd_cad_wbc_medications_ehr_analysis/src/main/.

## Availability of data and materials

Data from All of Us is publicly available. The Vanderbilt University Medical Center data that support the findings of this study are available from Vanderbilt University Medical Center but restrictions apply to the availability of these data, which were used under license for the current study, HIPPA protected, and so are not publicly available. Data are however available from the authors upon reasonable request and with permission of Vanderbilt University Medical Center. The data in question must first be reviewed by the Integrated Data Access and Services Core to ensure that the de-identification is complete and no potentially identifying information remains. Please contact the Vanderbilt Institute for Clinical and Translational Research for more information.

## Data Availability

Code and Data availability All code to conduct these analyses is available at https://bitbucket.org/davislabteam/mdd_cad_wbc_medications_ehr_analysis/src/main/. Availability of data and materials: Data from All of Us is publicly available. The Vanderbilt University Medical Center data that support the findings of this study are available from Vanderbilt University Medical Center but restrictions apply to the availability of these data, which were used under license for the current study, HIPPA protected, and so are not publicly available. Data are however available from the authors upon reasonable request and with permission of Vanderbilt University Medical Center. The data in question must first be reviewed by the Integrated Data Access and Services Core to ensure that the de-identification is complete and no potentially identifying information remains. Please contact the Vanderbilt Institute for Clinical and Translational Research for more information.

